# Monitoring maternal blood pressure variations in pregnancy: gestational age-specific reference percentiles and Z-scores

**DOI:** 10.1101/2025.09.04.25335123

**Authors:** Briana DeStaffan, Pauline Scherdel, Muriel Tafflet, Marie-Aline Charles, Vassilis Tsatsaris, Barbara Heude, Wen Lun Yuan

**Affiliations:** Université Paris Cité and Université Sorbonne Paris Nord, Inserm, INRAE, Centre for Research in Epidemiology and StatisticS (CRESS), F-75004, Paris, France; Unité Mixte Inserm-Ined-EFS ELFE, Ined 93300 Aubervilliers, France; Université Paris Cité, Department of Obstetrics and Gynecology, Port-Royal Maternity, Cochin Hospital, APHP, Paris, France

## Abstract

**Background:** Maternal blood pressure (BP) varies greatly during pregnancy in response to hemodynamic changes, which has led to debate surrounding the use of a single diagnostic threshold. Previous studies have generated “reference ranges” of BP in pregnancy, yet they lack implementation and translation to clinical practice. This study aimed to generate gestational age-specific references and analyze BP Z-scores to further explore cardiovascular dynamics in pregnancy and their potential clinical implications.

**Methods:** Repeated measurements of BP from 2 to 44 gestational weeks were extracted from the obstetric files of 1,875 mothers from the French EDEN cohort. Percentiles of systolic (SBP), diastolic (DBP), and mean arterial pressure (MAP) were modeled using Generalized Additive Models for Location, Scale, and Shape (GAMLSS) as a function of gestational age. They were generated from a “low-risk” reference population defined by reduced cardiovascular risk factors and no hypertensive disorders. For each woman in the overall sample, Z-scores of BP were calculated relative to the “low-risk” percentiles, to assess how BP deviated from the expected trajectory across gestation. BP Z-score trajectories according to hypertensive disorders of pregnancy categories (chronic hypertension, gestational hypertension, and preeclampsia) were then plotted and compared.

**Results:** A U-shaped trend was observed in overall and “low-risk” percentiles of SBP and MAP, with a nadir around 25 weeks and an increase in the last trimester. In the overall sample, SBP’s 95^th^ percentile curve remained below the 140mmHg diagnostic threshold between 15 and 35 weeks of gestation. The Z-score trajectories of women presenting with hypertensive disorders start to diverge as early as the first 5 gestational weeks, well-before their diagnoses (median age: 35-36 weeks). BP evolution differed according to type of hypertensive disorder, for example, a steep increase (>1 standard deviation) from early in pregnancy among preeclamptic women.

**Conclusion:** This percentiles-to-Z-score approach can position individual risk of mothers while considering the natural variation of blood pressure across pregnancy. Our results question the applicability of a non-time-specific threshold to these dynamics. Beyond their potential clinical applications, these references can be used in further research to examine “abnormal” cardiovascular trajectories and their consequences for future maternal and offspring health.

## Introduction

Hypertensive disorders of pregnancy (HDP) are among the leading causes of maternal and fetal morbidity and mortality around the world.^1^ The identification and monitoring of maternal blood pressure (BP) variations throughout pregnancy are crucial in improving HDP diagnoses from the first trimester,^2^ and for mitigating adverse outcomes such as fetal growth restriction, preterm birth, and long-term cardiometabolic consequences for both mother and child.^3–5^ Despite a growing body of literature on this subject, a refined understanding of BP fluctuations during pregnancy remains necessary to enhance risk stratification and early-stage intervention strategies.

During pregnancy, the maternal cardiovascular system undergoes substantial physiological adaptations, including increased vascular remodeling, elevated cardiac output, and plasma volume expansion, all of which contribute to variations in BP.^6^ Although BP evolution and changes across pregnancy have been well-documented,^6–8^ current clinical practice predominantly relies on fixed diagnostic thresholds for BP assessment.^9^ This approach may not fully capture the natural evolution of BP across gestation.

The International Society for the Study of Hypertension in Pregnancy (ISSHP) recommends routine BP monitoring as a standard component of prenatal care.^9^ However, current practices rely largely on absolute BP values (mmHg), irrespective of gestational age. Growing evidence suggests that implementing gestational age-specific BP percentiles in routine obstetric care could improve the identification of abnormal BP deviations from physiological norms.^10–12^

Using obstetric records from the EDEN French birth cohort, this study presents the development of internal, gestational age-specific percentiles of maternal blood pressure in pregnancy. To provide evidence of their potential utility, we illustrate how BP Z-scores derived from these percentiles can discriminate divergent and potentially adverse blood pressure trajectories during pregnancy.

## Methods

This work is reported following the Strengthening the Reporting of Observational Studies in Epidemiology (STROBE) guidelines (**S1**, Strobe Checklist).^13^

### Study population

The data used in this study were sourced from the EDEN mother-child cohort, which is described in detail elsewhere.^14^ From 2003 to 2006, two university hospitals in the French cities of Nancy and Poitiers enrolled 2,002 pregnant women before their 24^th^ week of gestation. Exclusion criteria for recruitment consisted of maternal diabetes before pregnancy, multiple pregnancy, intention to deliver outside of the study hospitals or move out of the region in the following 3 years, and inability to speak French. The EDEN study was approved by the ethics research committee of Kremlin-Bicêtre hospital (ID 0270, December 12, 2002) and by the French Data Protection Authority (CNIL, ID 902267, December 12, 2002). Upon inclusion, each mother provided written informed consent for participation. From the 2,002 women initially included in the EDEN cohort, 1,875 had an available obstetric record containing at least 1 blood pressure measurement (systolic-diastolic pair).

### Maternal blood pressure measurements

Longitudinal measurements of systolic (SBP) and diastolic (DBP) BP (mmHg) were extracted from obstetric files with a maximum of 12 measurements (median (IQR) = 8 (6-9)), ranging from 2 to 44 gestational weeks. Some measurements of DBP <20mmHg were recorded (n=5 measurements). These were deemed clinically-implausible and removed from the dataset. Mean arterial pressure (MAP) was calculated using the following formula: diastolic BP + 1/3 (systolic BP – diastolic BP).^15^

### Maternal Characteristics

Gestational age was calculated as the difference between date of delivery and last menstrual period (declared by the participants or, if unavailable, extracted from their obstetric files). Mothers’ obstetric records included information on diagnoses of hypertensive disorders of pregnancy, including pre-existing (chronic) hypertension, gestational hypertension, and preeclampsia, and/or any history of these disorders in a previous pregnancy. Gestational hypertension is defined as an SBP ≥ 140mmHg and/or a DBP ≥ 90mmHg after 20 weeks’ gestation, in the absence of pre-existing hypertension. Preeclampsia is defined by the same features along with the presence of proteinuria.^9^ Preeclampsia was also categorized into early-onset and late-onset, at <34 and ≥34 weeks of gestation, respectively.^16^ Diagnoses of gestational diabetes were also extracted from obstetric records.

During a face-to-face interview at 24-28 weeks of gestation, mothers reported their country of birth, level of education (years of education), monthly household income, and pre-pregnancy weight. Tobacco use (number of cigarettes per week) during pregnancy was also evaluated at 24-28 weeks, and at an additional interview at delivery. Smoking status was consolidated into a binary variable: whether the mother smoked during pregnancy or not. Pre-pregnancy body mass index (BMI) was derived using measured height at the cohort’s baseline clinical examination at 24-28 weeks of gestation and self-reported pre-pregnancy weight.

### Neonatal Covariates

Birthweight Z-scores have been previously generated in the EDEN cohort based on a customized fetal growth curve built for French children, integrating maternal height and pre-pregnancy weight, parity, offspring’s sex, and gestational age.^17,18^ Newborns with birthweight Z-scores below the 10^th^ percentile were classified as small-for-gestational age (SGA). Conversely, newborns with birthweight Z-scores over the 90^th^ percentile were classified as large-for-gestational age (LGA). Prematurity was defined as delivery <37 weeks of gestation.

### “Low-risk” subsample

From the overall sample of 1,875 with at least one BP measurement available, a “low-risk” sample (n = 795) was defined, purported to be at lower cardiovascular risk, i.e. with none of the recognized HDP risk factors and without HDP. The inclusion criteria for this internal reference sample were as follows: no chronic hypertension, gestational hypertension, or preeclampsia; no history of hypertensive disorders of pregnancy; no gestational diabetes; BMI < 30 kg/m2; age at delivery <40 years; no smoking during pregnancy; no preterm delivery; no offspring born small- or large-for gestational age. These criteria were defined in alignment with recognized HDP risk factors and previous literature that has generated reference ranges of BP during pregnancy.^10–12^

Due to the low percentage of missing data for each covariate (**Table 1**) and to optimize the specificity of the “low-risk” sample, individuals with missing data for any of the inclusion criteria were excluded from this sample.

**Table 1.**
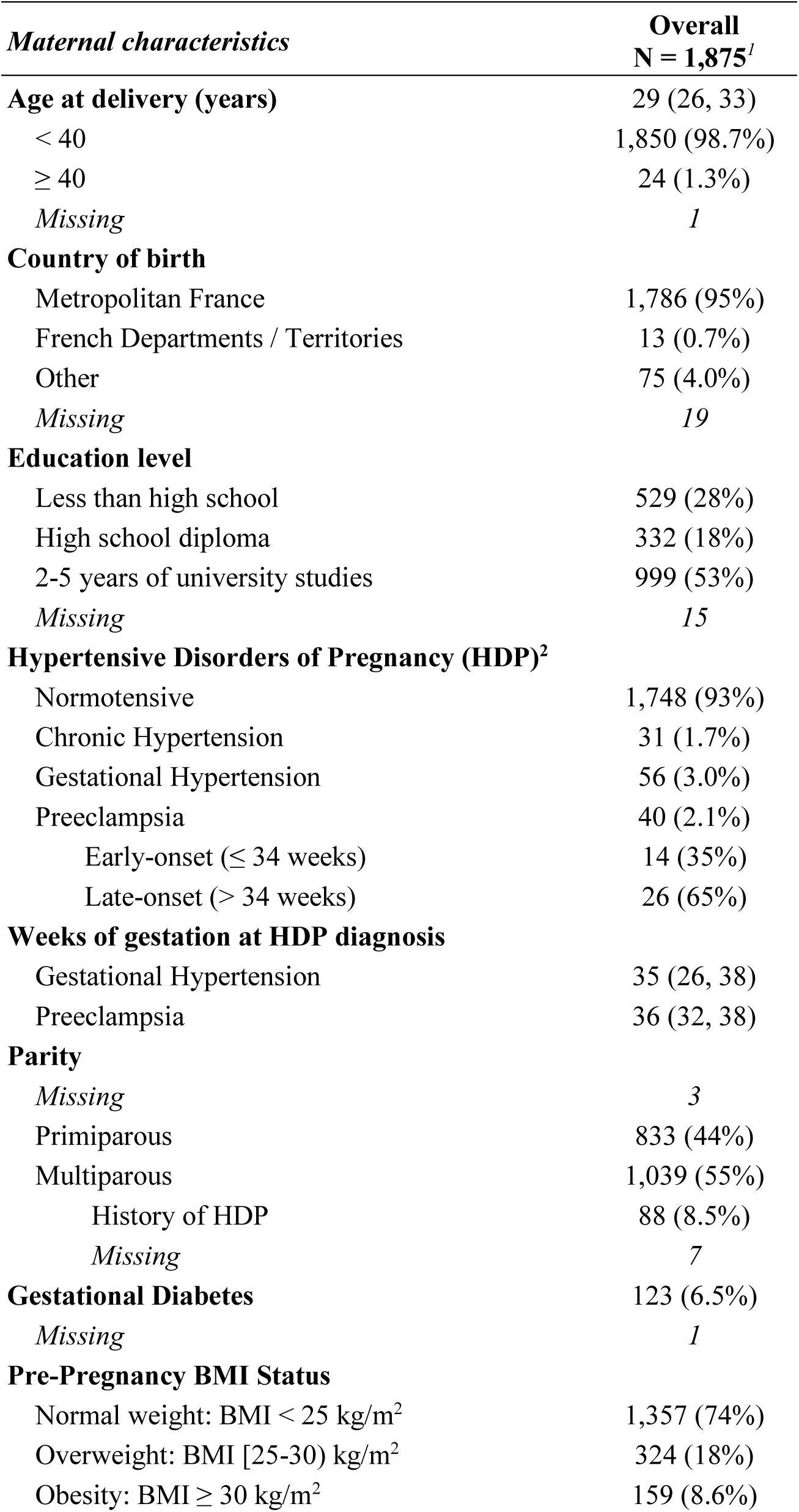
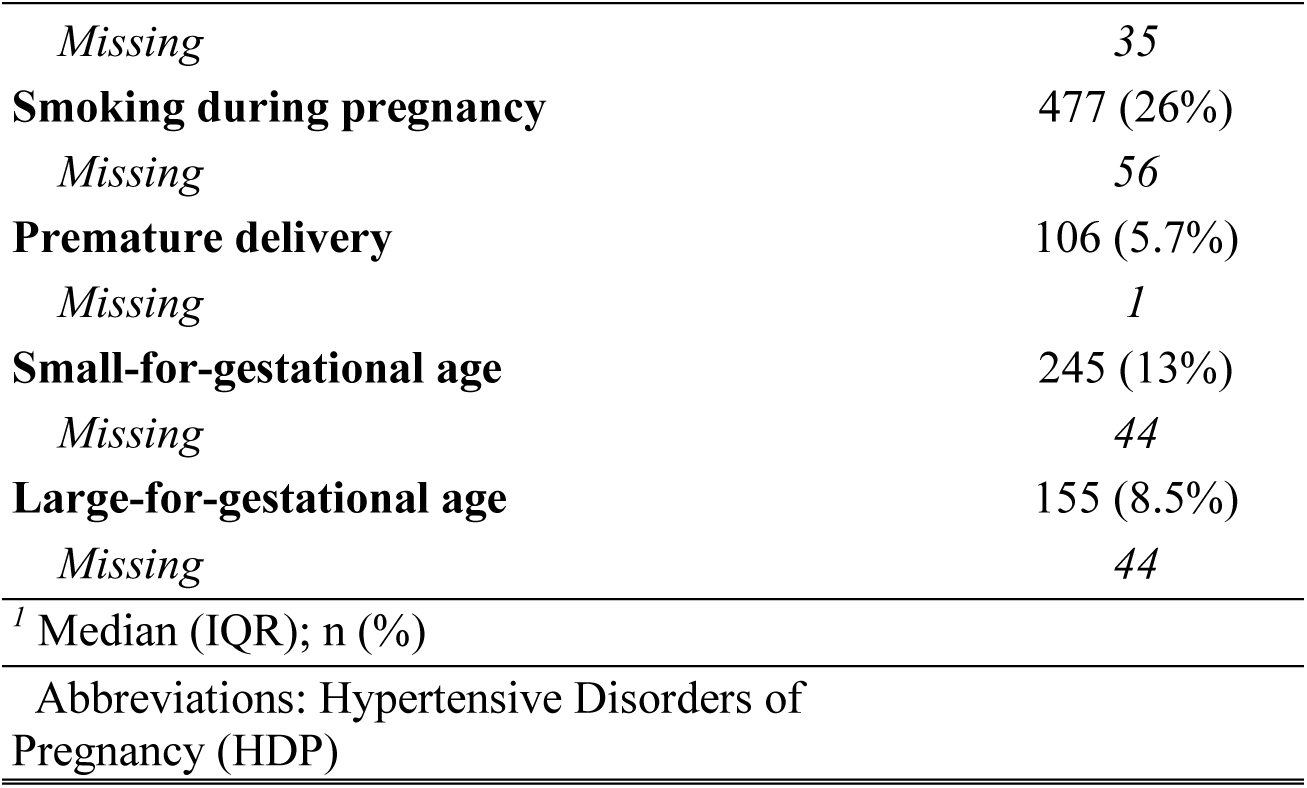
Population description.

| *Maternal characteristics* | **Overall**<br>**N = 1,875<sup>1</sup>** |
| --- | --- |
| **Age at delivery (years)** | 29 (26, 33) |
| < 40 | 1,850 (98.7%) |
| ≥ 40 | 24 (1.3%) |
| *Missing* | 1 |
| **Country of birth** |  |
| Metropolitan France | 1,786 (95%) |
| French Departments / Territories | 13 (0.7%) |
| Other | 75 (4.0%) |
| *Missing* | 19 |
| **Education level** |  |
| Less than high school | 529 (28%) |
| High school diploma | 332 (18%) |
| 2-5 years of university studies | 999 (53%) |
| *Missing* | 15 |
| **Hypertensive Disorders of Pregnancy (HDP)<sup>2</sup>** |  |
| Normotensive | 1,748 (93%) |
| Chronic Hypertension | 31 (1.7%) |
| Gestational Hypertension | 56 (3.0%) |
| Preeclampsia | 40 (2.1%) |
| Early-onset (≤ 34 weeks) | 14 (35%) |
| Late-onset (> 34 weeks) | 26 (65%) |
| **Weeks of gestation at HDP diagnosis** |  |
| Gestational Hypertension | 35 (26, 38) |
| Preeclampsia | 36 (32, 38) |
| **Parity** |  |
| *Missing* | 3 |
| Primiparous | 833 (44%) |
| Multiparous | 1,039 (55%) |
| History of HDP | 88 (8.5%) |
| *Missing* | 7 |
| **Gestational Diabetes** | 123 (6.5%) |
| *Missing* | 1 |
| **Pre-Pregnancy BMI Status** |  |
| Normal weight: BMI < 25 kg/m<sup>2</sup> | 1,357 (74%) |
| Overweight: BMI [25-30) kg/m<sup>2</sup> | 324 (18%) |
| Obesity: BMI ≥ 30 kg/m<sup>2</sup> | 159 (8.6%) |

**Table 1. Population description**
| *Maternal characteristics* | **Overall**<br>**N = 1,875<sup>1</sup>** |
| --- | --- |
| *Missing* | 35 |
| **Smoking during pregnancy** | 477 (26%) |
| *Missing* | 56 |
| **Premature delivery** | 106 (5.7%) |
| *Missing* | 1 |
| **Small-for-gestational age** | 245 (13%) |
| *Missing* | 44 |
| **Large-for-gestational age** | 155 (8.5%) |
| *Missing* | 44 |
| <sup>1</sup> Median (IQR); n (%) |  |
| Abbreviations: Hypertensive Disorders of<br>Pregnancy (HDP) |  |

### Statistical analyses

#### Gestational age-specific percentiles of blood pressure

Percentile curves of maternal SBP, DBP, and MAP across gestational age were generated using the Generalized Additive Models for Location, Scale, and Shape (GAMLSS) method and corresponding statistical package in R.^19^ All statistical analyses were performed in R version 4.2.1. After testing multiple available distributions, we chose the box-cox power estimation (BCPE) distribution for both its applicability to our data and its comparability with pre-existing literature.^10^ Further explanations of model specification, fit, and selection are detailed in **Supplemental Methods (S2)**.

Percentile curves of SBP, DBP, and MAP across gestation were calculated in the “low-risk” sample. Percentiles were also calculated in the overall sample, in a supplementary analysis, for descriptive and comparative purposes. We tested the internal validity of the “low-risk” reference percentiles using an internal empirical evaluation by manually calculating median BP in the “low-risk” sample in 5-week increments, and overlaying these values with the 50^th^ percentile (median) estimated by the models.

The “low-risk” reference percentiles estimate three key parameters for each day of gestation: L (lambda), representing the Box-Cox power transformation for skewness, M (mu), the median, and S (sigma), the coefficient of variation. Together, these parameters describe the distribution of a given measure (maternal BP) across time (gestational age). From our models of SBP, DBP and MAP, we extracted “low-risk” reference estimations for L, M, and S for each gestational age (expressed in days).

#### Gestational age-specific blood pressure Z-scores

The LMS values enable the calculation of gestational age-specific Z-scores, which standardize individual BP measurements relative to the “low-risk” reference distribution. Each Z-score is computed using the following equation (**Equation 1**):^20^

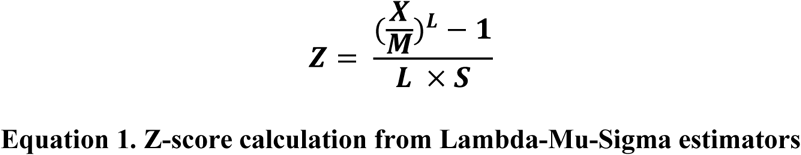

where ***X*** is the observed measurement (maternal BP in mmHg), and ***L, M***, and ***S*** correspond to the estimated values from the model for the specific gestational age at which ***X*** was measured.

This transformation ensures that the Z-scores are 1) gestational age-specific, accounting for expected BP changes throughout pregnancy, and 2) adjusted for distributional characteristics, incorporating variability in the median, variance, and skewness observed in the “low-risk” sample across gestational age.

Based on these internal references, gestational age-specific BP Z-scores were calculated for all women in the study sample (N=1,875).

#### Trajectories of blood pressure Z-scores

To evaluate BP’s physiology in our sample, we plotted trajectories of maternal BP Z-scores across gestation using smoothing splines. We first plotted BP Z-scores corresponding to the recommended hypertensive thresholds of 140mmHg and 90mmHg.^9^

Then, we plotted trajectories of BP Z-scores across gestation, stratified by the categories of HDP: normotensive, chronic hypertension, gestational hypertension, and preeclampsia. Any women in the “low-risk” sample were excluded from the Normotensive category, as their Z-scores would bias the overall trajectory towards 0. If women had any BP measurements after their HDP diagnosis, these data were excluded from the trajectories to limit the potential bias of intervention, treatment, or enhanced surveillance.

## Results

### Sample characteristics

Characteristics of the overall sample are described in **Table 1**. Mothers had a median age of 29. The prevalence of hypertensive disorders of pregnancy (incl. chronic hypertension, gestational hypertension, and preeclampsia) was 6.7%. Median gestational ages at diagnosis of gestational hypertension and preeclampsia were 35 and 36 weeks of gestation, respectively. Among mothers with preeclampsia, 35% were early-onset and 65% were late-onset. Demographic characteristics of the “low-risk” sample, as compared to the overall, are available in **Supplementary Table 1 (S3).**

### Gestational age-specific BP percentiles: Overall and “low-risk”

Gestational age-specific percentiles in the “low-risk” (N = 795) sample are illustrated in **Figure 1A** (SBP) and **Figure 2A** (DBP), respectively. Percentiles for MAP are included in **Supplemental Figure 1 (S4)**. Corresponding reference (LMS) values for SBP, DBP, and MAP are each available in **Supplementary Table 2 (S5).** For comparison purposes, percentiles generated in the overall sample for SBP and DBP are included in **Supplemental Figure 2 (S6).**

**Figure 1.**
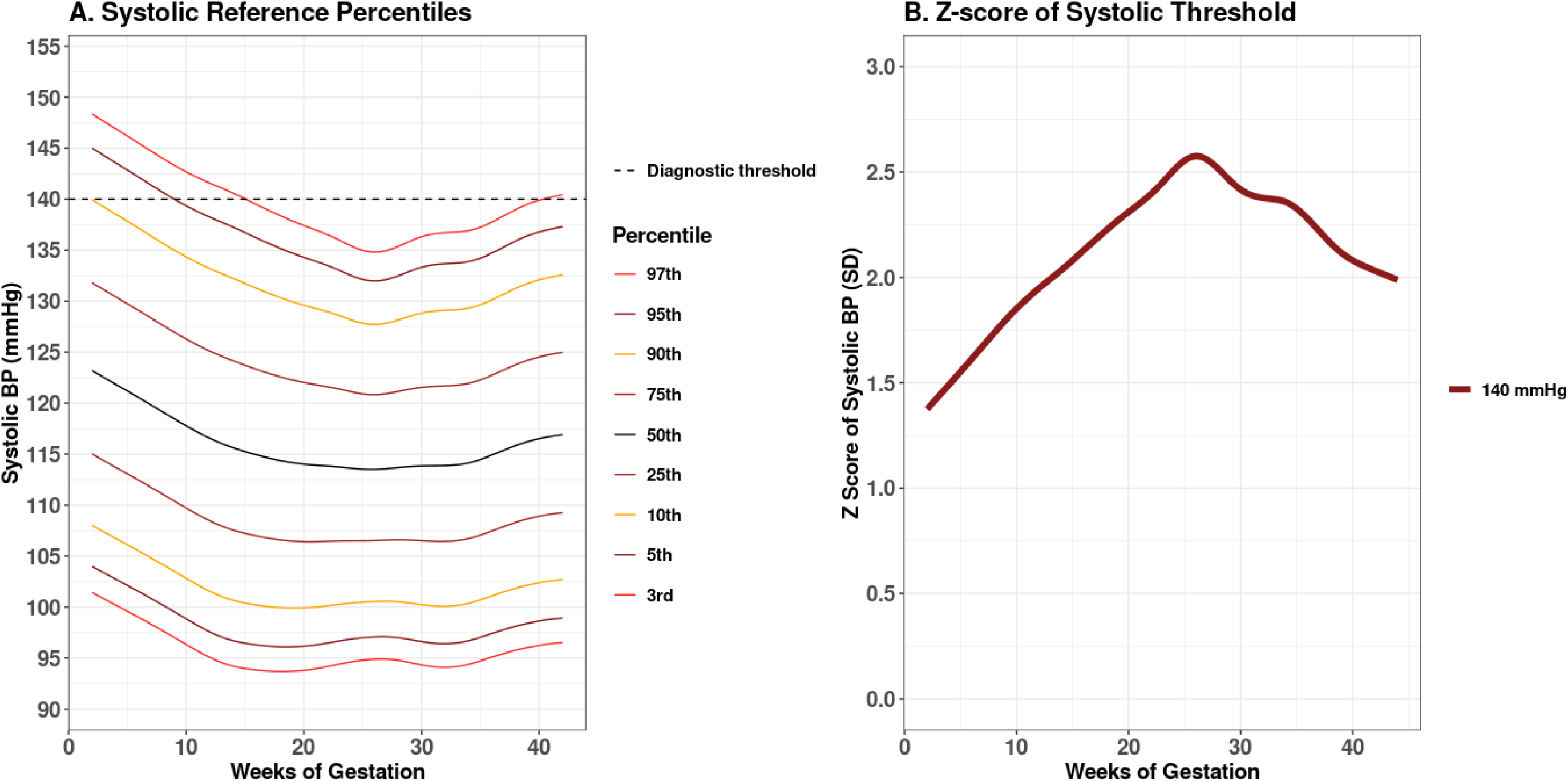
A. Gestational age-specific reference percentiles of maternal systolic blood pressure (BP) across pregnancy in the EDEN cohort, from the “low-risk” sample (N=795). B. Gestational age-specific maternal BP Z-scores of the 140mmHg systolic hypertensive threshold.

**Figure 2.**
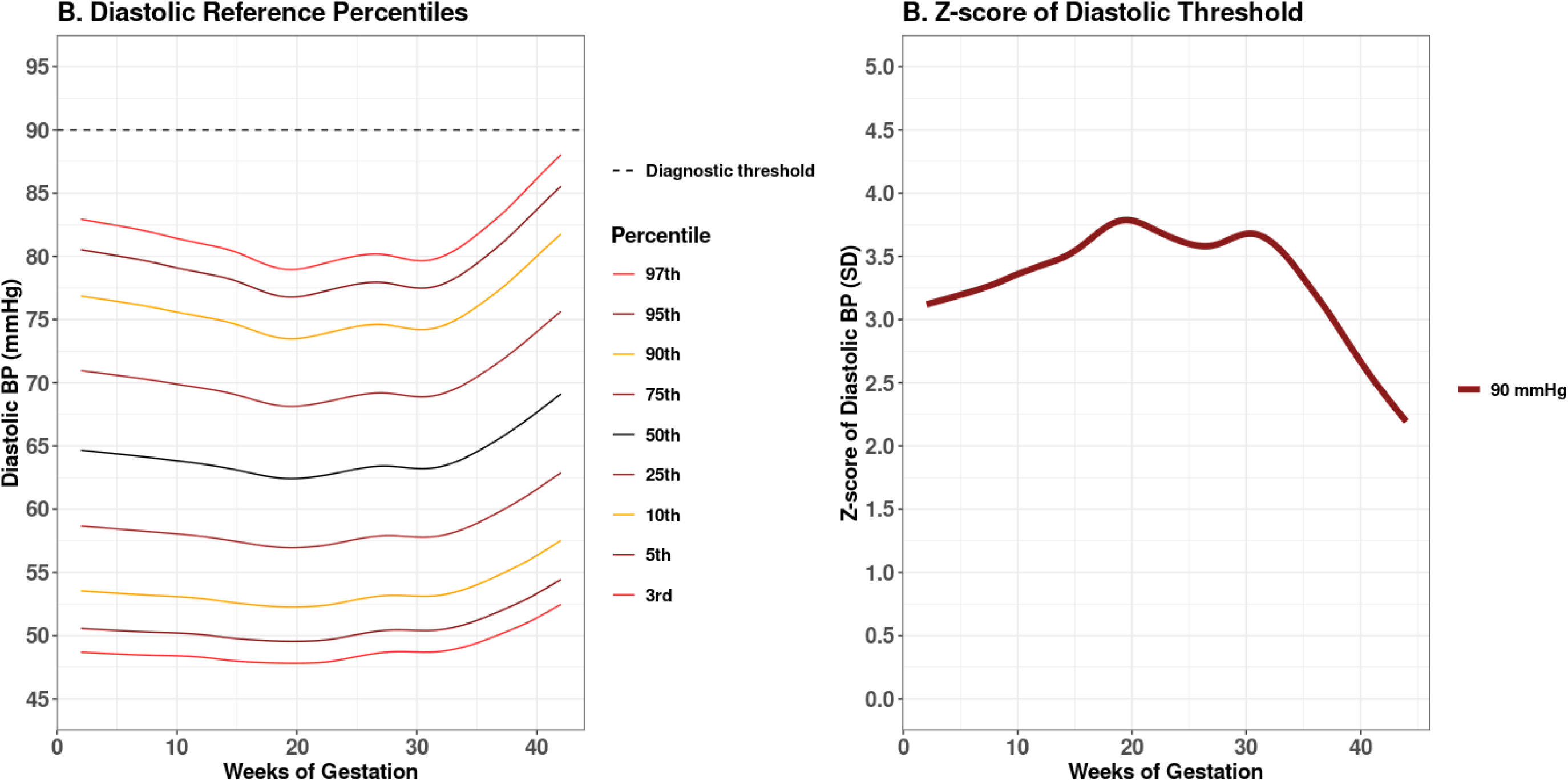
A. Gestational age-specific reference percentiles of maternal diastolic blood pressure (BP) across pregnancy in the EDEN cohort, from the “low-risk” sample (N=795). B. Gestational age-specific maternal BP Z-scores of the 90mmHg diastolic hypertensive threshold.

The overall BP patterns between the overall and “low-risk” samples were similar. As expected, after excluding mothers with hypertensive disorders and risk factors, the “low-risk” reference ranges shifted to reflect lower blood pressure values compared to the overall sample.

The median (50^th^ percentile) SBP in the “low-risk” sample exhibited a U-shaped pattern, decreasing from 121mmHg at 5 weeks to its lowest value of 114mmHg around mid-gestation (26 weeks), then gradually rose to 117mmHg by 44 weeks.

Higher SBP percentiles showed a greater decline in SBP to reach their nadirs. Lower percentiles (<25^th^) still descended and rose across gestation, yet did not display the same U-shaped trend, indicating asymmetry in SBP changes across the distribution.

MAP percentile patterns closely mirrored those of SBP, with a similar mid-gestation decline observed across percentiles. In contrast, DBP trends differed. In the “low-risk” group, median DBP reached its lowest value earlier, at 19 weeks, after a modest descent of 3mmHg. It then remained relatively stable before increasing in late pregnancy.

In the internal empirical evaluation, illustrated in **Supplemental Figure 3 (S7)**, the median percentiles estimated for SBP and DBP generally tracked with the observed medians of SBP and DBP in the “low-risk” population. However, the median percentiles of MAP demonstrated a better alignment with its observed median at each gestational age.

### Interpreting the clinical BP thresholds for hypertension across gestation

#### Systolic BP

In the “low-risk” sample, the 95^th^ SBP percentile remained above the clinical threshold for hypertension (140mmHg) until approximately 10 weeks’ gestation, after which it declined below this cutoff (**Figure 1A**). In contrast, the 90^th^ SBP percentile remained below 140mmHg from as early as 5 weeks, in both samples. In the overall sample, the 95^th^ SBP percentile also dropped below 140mmHg between 15 and 35 weeks.

When the 140mmHg threshold is expressed as a gestational age-specific Z-score of SBP, its value varies greatly across time, particularly during mid-gestation, where even the highest SBP percentiles are the furthest from the threshold (**Figure 1B**). At 5 weeks of gestation, an SBP of 140 mmHg corresponds to a Z-score of 1.6 SD but increases to 2.6 SD at the expected nadir at 26 weeks (a 1 SD increase). At 35 weeks – the median gestational age for diagnosis of gestational hypertension in this sample – a SBP of 140 mmHg corresponds to a Z-score of 2.3 SD, decreasing to 2.0 SD by 42 weeks.

#### Diastolic BP

In both the overall and “low-risk” samples, even the highest percentiles of DBP did not cross the 90mmHg diagnostic threshold. The 95^th^ percentile of DBP declined from 80mmHg at 5 weeks to 77mmHg at 19 weeks, and increased to 86mmHg at 44 weeks (**Figure 2A**). DBP remained stable across all percentiles at 26 weeks, which is the gestational age at nadir for both SBP and MAP.

When expressed as a gestational age-specific Z-score, a DBP of 90mmHg corresponds to higher Z-scores than those of SBP, peaking at 3.8 SD at 19 weeks (the observed nadir for DBP in this sample), and decreasing to 2.4 SD by 42 weeks (**Figure 2B**).

### Gestational age-specific BP Z-score trajectories in HDP

Gestational age-specific BP Z-score trajectories varied between women with and without hypertensive disorders of pregnancy (HDP). In **Figure 3**, we can observe divergences in SBP Z-scores (which represent deviations from the “low-risk” reference curve), as early as before 10 weeks, in women who later developed HDP. These deviations were apparent well before the median gestational age at diagnosis (35–36 weeks).

**Figure 3.**
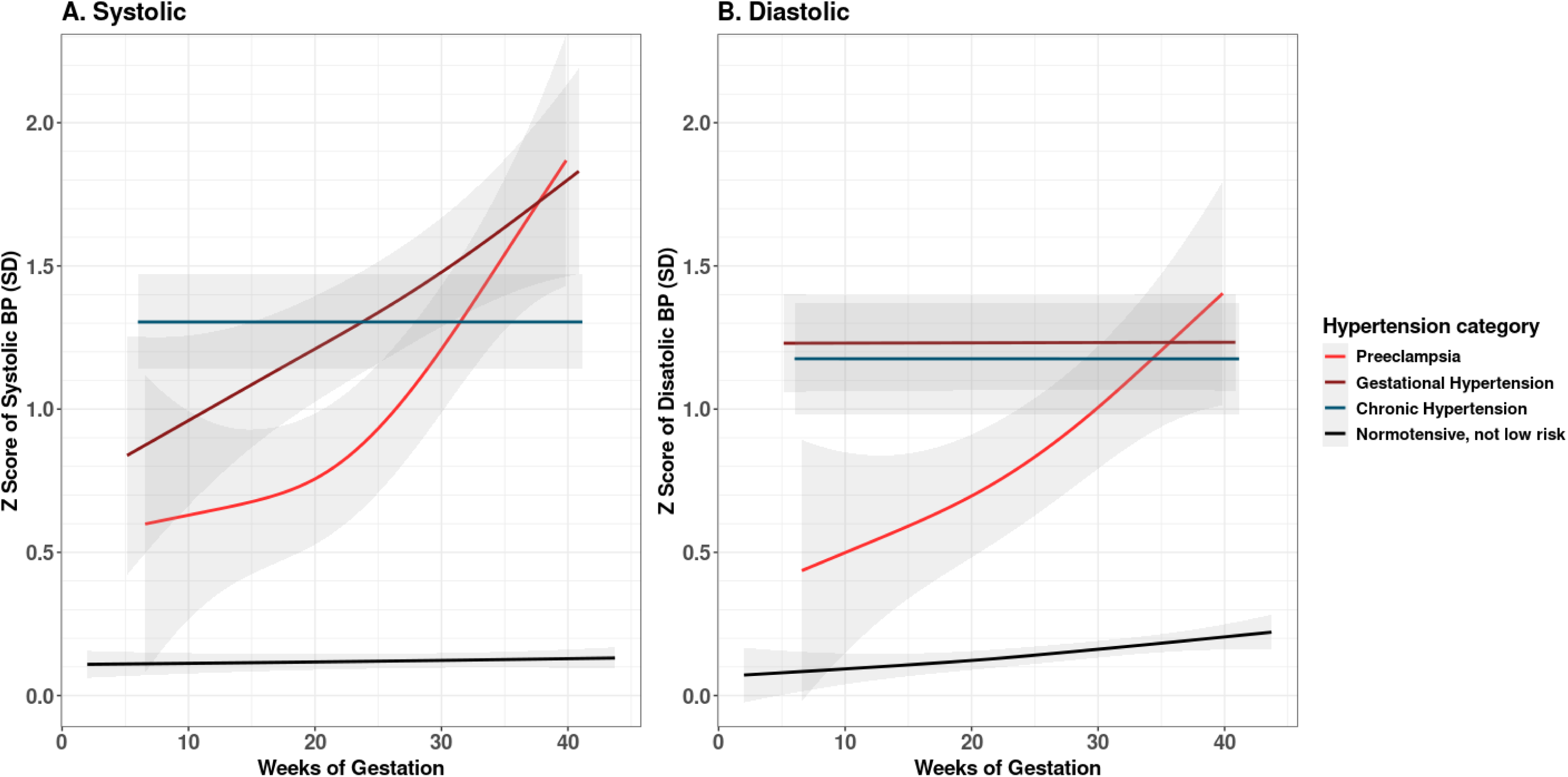
Gestational age-specific Z-scores of maternal blood pressure across pregnancy in the EDEN cohort, according to hypertensive disorders of pregnancy categories. Systolic and diastolic Z-score trajectories and 95% confidence intervals are illustrated in panels A and B, respectively. Data from normotensive women who were included in the “low-risk” reference subsample are excluded.

Patterns of deviation differed according to the type of hypertensive disorder. As expected, normotensive mothers (excluding those in the low-risk sample) exhibited a stable trajectory, with SBP Z-scores remaining close to 0 throughout pregnancy. Mothers with chronic hypertension had higher initial SBP Z-scores than normotensive mothers, yet maintained a stable trajectory throughout pregnancy. In contrast, mothers who developed gestational hypertension initially had lower SBP Z-scores than those with chronic hypertension, but experienced a steady increase across gestation until time of diagnosis. Women who developed preeclampsia also exhibited a sustained and steep increase in SBP Z-scores, with values rising by almost 1.5 standard deviations across pregnancy. This trajectory also began at a lower level than that of gestational hypertension.

DBP Z-score trajectories in normotensive and pre-eclamptic women followed the trends of their SBP Z-scores. However, in contrast to their SBP trajectories, those with gestational hypertension exhibited a similar trend to that of chronic hypertension, maintaining a stable, yet elevated, level of DBP Z-scores across gestation. All MAP Z-score trajectories of HDP followed the trends of their corresponding SBP Z-score trajectories (**Supplemental Figure 4, S8**).

## Discussion

This study aimed to address the wide variation in maternal BP during pregnancy by generating gestational age-specific reference percentiles in a general population. When examining changes in corresponding gestational age-specific BP Z-scores among women who later developed HDP, deviations from normotensive women were observed as early as before 10 weeks of gestation. The trajectory of BP Z-scores varied according to the type of HDP, with distinct patterns observed between gestational hypertension and preeclampsia. The threshold to diagnose gestational hypertension of 140/90mmHg, currently used in clinical practice, corresponded to different SBP and DBP Z-scores across gestational age, indicating that one, fixed threshold might not best reflect a dynamic phenomenon. To our knowledge, this is the first study to calculate clinically-interpretable gestational age-specific BP Z-scores, derived from reference percentiles, providing a novel tool for future research to better-understand BP dynamics in pregnancy and associated factors.

In the maternal BP percentiles, we observed a mid-pregnancy drop in SBP and MAP which aligns with established physiological patterns.^7,21–23^ A recent publication using the same method has contested the existence of this nadir in blood pressure.^24^ This discrepancy could be explained by the gestational age period covered (often starting at 10 or 12 weeks). As our study includes data as early as 2 weeks of gestation, the availability of data from very early in pregnancy might allow for our models to capture a potential descending trend.

The internal empirical evaluation, conducted to assess internal validity, demonstrated that the median estimated in the MAP reference percentiles remained close to the observed median MAP values in the “low-risk” population across gestation. This supports the reliability of the MAP model in capturing central tendencies over time. However, the SBP and DBP reference percentiles diverged slightly from their observed medians, particularly after 30 weeks of gestation. Despite these differences, diagnostic assessments (see Supplemental Methods) indicated that the SBP and DBP models maintained an adequate overall fit, capturing the distributional shape of SBP and DBP across gestation. One plausible explanation for the difference between estimated and observed medians lies in a phenomenon inherent to clinical practice, end-digit preference.^25^ Our data demonstrate a tendency towards rounded values (i.e. a DBP of 60, 70, 80 mmHg, etc.). Therefore, although a high concentration of datapoints exists around the observed DBP median of 60mmHg at 30 weeks, for example, the estimated median may be closer to the high concentrations of points also at 70 and 80 mmHg. The estimated median is still effectively describing an overall trend in the data’s distribution, but is biased away from the observed median by the multimodal nature of the rounded values. In any case, the percentiles-based approach enables the smoothing of potential bias in observed measurements due to end-digit preference.

In contrast, the MAP reference percentiles demonstrated strong empirical alignment with the observed MAP medians. This suggests that MAP may offer a more robust and stable measure of central BP tendency in pregnancy, potentially making it more suitable for reference standards and clinical assessment. Particularly in the context of HDP, a systematic review and meta-analysis has previously evaluated MAP as a better predictor of later preeclampsia than SBP or DBP alone.^26^ Given that MAP integrates both SBP and DBP, it may mitigate some of the variability or rounding observed in DBP or SBP alone, by leveraging the behavior of the other over time.

The SBP deviation trajectories that we observed in different HDP categories are consistent with previous studies of BP changes during pregnancy.^7,8,23^ Our findings suggest that deviations in BP trajectories arise early in pregnancy and may follow trends specific to certain hypertensive disorders. These distinct patterns, previously suggested in the literature, are more clearly delineated in our analysis using gestational age-specific Z-scores.

However, it is important to contextualize the interpretation of these trajectories. By design, for these illustrations, we excluded all BP measurements taken after HDP diagnosis, to avoid the bias of treatment effects or post-diagnosis interventions. While this approach improves the validity of BP trajectories leading up to clinical intervention, it also introduces a degree of uncertainty regarding the shape and interpretation of group-level trends according to gestational age at diagnosis. For instance, the smoothed trajectories for gestational hypertension include individuals diagnosed both shortly after 20 weeks and those diagnosed much later in pregnancy. This averaging may obscure distinct inflection points or progression rates specific to the timing of hypertension onset.

The preeclampsia trajectory, however, demonstrates a more sustained and steeper rise in SBP, DBP, and MAP Z-scores, with the most prominent inflection around the mid-pregnancy nadir (26 weeks). This increase, which follows a different shape than that of gestational hypertension, may reflect a distinct pathophysiological process. These different trajectory shapes lend further support to the notion that different HDP phenotypes may have unique etiological pathways and gestational age-specific dynamics.^7^ In fact, an emerging body of literature has illustrated the clinical relevance of maternal BP trends in early pregnancy. Gunderson and colleagues estimated BP trajectories before 20 weeks’ gestation and found that increasing BP trends during this period could predict HDP later in pregnancy.^2^ Our work proposes the additive value of using gestational age-specific Z-scores to surveille how BP changes, relative to its distribution at each week of gestation, in a deeper investigation of these trends.

A key observation from our results is that the upper percentiles of SBP fell below the conventional hypertensive diagnostic threshold (140mmHg) during a substantial portion of gestation. In the overall, non-restricted sample, the 95^th^ percentile of SBP remained below this threshold between 15 and 35 weeks of gestation. Notably, this period precedes the median gestational age of diagnosis of HDP in our cohort, which occurred between 35 and 36 weeks.

This “mismatch” in timing highlights the methodological limitation of applying a static diagnostic threshold to the entire pregnancy period. When 140mmHg is expressed as a gestational age-specific Z-score, according to our “low-risk” reference distribution, its standardized value varies considerably, according to gestational age. Otherwise said, the same absolute value of 140mmHg represents very different degrees of deviation from the “expected” BP norm, at different gestational ages.

These findings underscore the dynamic nature of maternal BP during pregnancy, particularly SBP, and support existing concerns regarding the adequacy of static diagnostic criteria.^27,28^ Several studies have proposed lowering the SBP threshold to 130mmHg to improve early detection, but our results suggest that a more fundamental shift in approach may be warranted. Specifically, dynamic, gestational age-adjusted thresholds – or continuous risk models incorporating Z-scores – may better account for physiological variation of BP in pregnancy and improve both surveillance and risk stratification during the prenatal period.

### Strengths and limitations

This study is strengthened by the nature of our data - repeated BP measurements throughout pregnancy - which have the advantage of longitudinal follow-up and add dimension to our analyses. These data are available from very early in pregnancy (even before 10 gestational weeks), which is less-represented in the available literature. In addition to the BP data, we have comprehensive patient-reported and clinical data available during pregnancy, due to the nature of the cohort’s design. Finally, to our knowledge, this study is the first of its kind to evaluate BP diagnostic thresholds in relation to gestational age and analyze BP Z-score trajectories derived from gestational age-specific references. This Z-score approach has the potential for extensive future use in research and clinical settings.

This study has some limitations. The EDEN cohort is highly educated, and over 95% of mothers were born in France. The BP data used were retrospectively extracted from obstetric records, and measurements were conducted according to each hospital’s standard operating procedures. As such, variation in measurement conditions (e.g., device type, patient posture, timing) may have introduced heterogeneity and measurement error. This could, for example, help explain why even the upper DBP percentiles remained below the diagnostic threshold throughout pregnancy. On the other hand, this variation reflects how BP monitoring and diagnoses are made in everyday clinical settings. The resulting reference ranges and derived Z-scores are specific to the EDEN cohort and may not be generalizable to more diverse populations.

This highlights the need for external validation and future replication of our findings in other settings to assess their broader applicability. Future work analyzing BP Z-scores could benefit from stratifying by gestational age of HDP diagnosis, severity, and other maternal characteristics. Nonetheless, this study represents a necessary first step towards more individualized, physiologically-relevant, and interpretable blood pressure monitoring in pregnancy.

### Clinical Implications

Indeed, this study opens perspectives to a wide variety of future research perspectives and exploration of the potential impact of applying these references in clinical practice. Gestational age-specific BP Z-scores can be used, for example, to define an abnormal BP, or to evaluate the predictive ability of BP deviation for future HDP or maternal cardiovascular disorders later in life. Health determinants, clinical complications, and adverse maternal and fetal outcomes have been widely studied with raw BP trajectories, and would merit replication with BP Z-scores to evaluate how the consideration of variation according to gestational age might impact, contradict, or enhance these results. From a clinical perspective, translating BP Z-scores into clinical practice will require additional steps, including user-friendly tools, clinician training, and integration into electronic health record systems. Beyond the individual level, maternal BP Z-scores introduce a promising complement to raw BP measurements in pregnancy follow-up that could ultimately aid in strengthened clinical approaches and public health initiatives to improve pregnancy health.

## Conclusions

Gestational age-specific references and corresponding Z-scores allow for a standardized assessment of maternal BP through comparison to a reference population. This approach accounts for the inherent variability of BP in pregnancy, facilitating early detection of deviations from expected physiological patterns. External evaluation of the references and the Z-score trajectories is a critical next step, to compare and assess their validity in diverse populations, particularly to confirm their utility in identifying and characterizing women who develop HDP. The use of Z-scores enables individualized risk assessment that is specific to gestational age. This can enhance future research to improve BP surveillance and potentially facilitate earlier management of trajectories at higher risk, especially when deviations occur before crossing the diagnostic threshold.

Further research should assess whether BP Z-score thresholds can outperform raw BP values in the prediction of HDP. It is important to investigate whether Z-score-based monitoring could add additional clinical value to current practice. The development and validation of evidence-based Z-score thresholds could inform personalized surveillance strategies and support clinical decision-making, in the interest of early intervention.

## Supporting information

STROBE Checklist

## Data Availability

The data underlying the findings cannot be made freely available for ethical and legal restrictions imposed, due to a substantial number of variables which, together, could be used to re-identify participants. However, data and analytic code can be obtained upon request from the EDEN steering committee (contact).

## Acknowledgments

We express our deep gratitude to the mothers and families who have participated in the EDEN cohort and continue to volunteer their valuable time and energy in helping further this research. The authors thank the EDEN mother–child cohort study group, whose members are I Annesi-Maesano, JY Bernard, J Botton, M-A Charles, P Dargent-Molina, B de Lauzon-Guillain, P Ducimetière, M de Agostini, B Foliguet, A Forhan, X Fritel, A Germa, V Goua, R Hankard, B Heude, M Kaminski, B Larroque, N Lelong, J Lepeule, G Magnin, L Marchand, C Nabet, F Pierre, R Slama, MJ Saurel-Cubizolles, M Schweitzer, O Thiebaugeorges.

## Non-Standard Abbreviations List

HDP: Hypertensive disorders of pregnancy
GAMLSS: Generalized Additive Models for Location, Scale, and Shape

