## Supplemental Figures for "Monitoring maternal blood pressure variations in pregnancy: gestational age-specific reference percentiles and Z-scores"

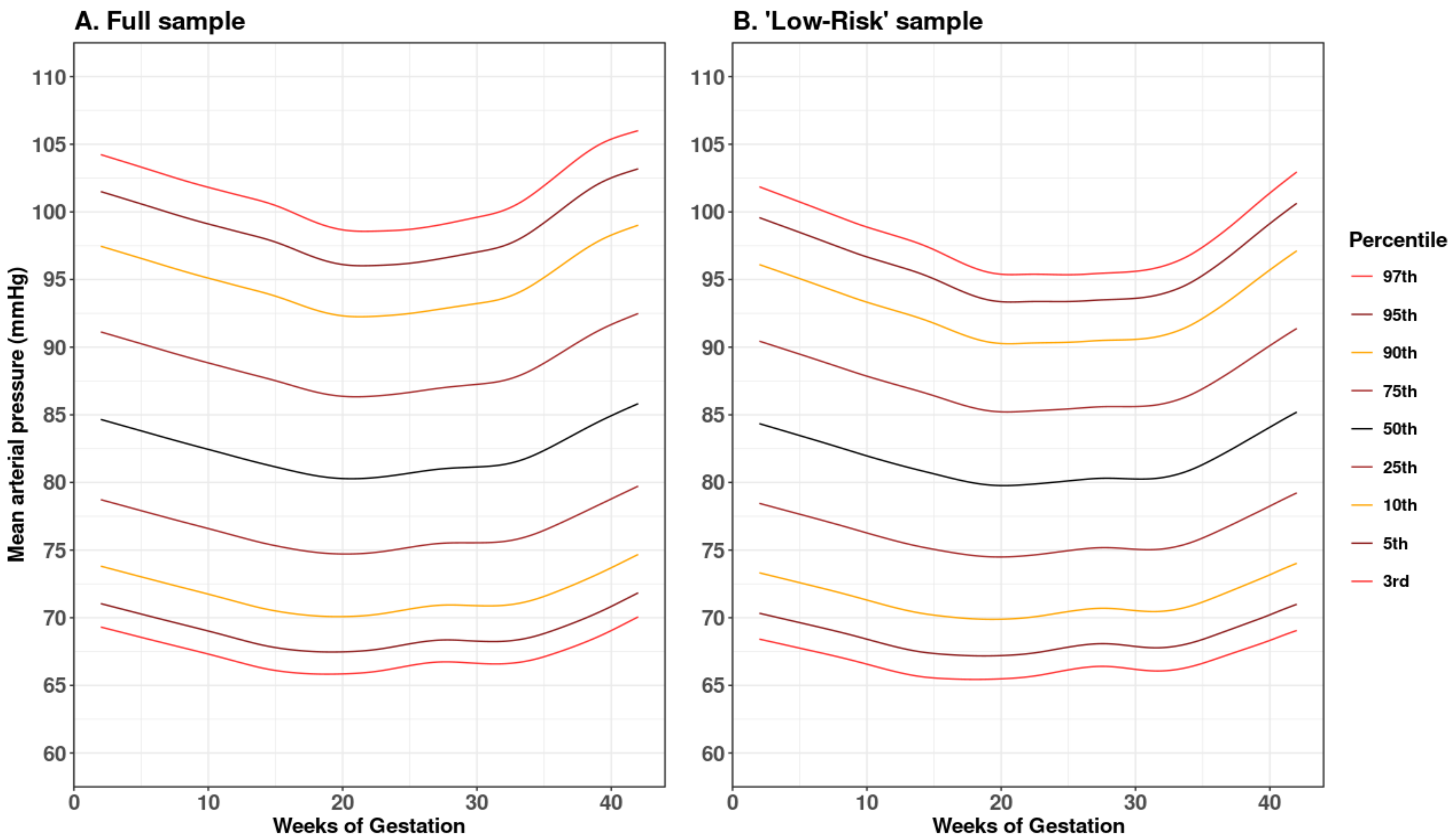


**Supplemental Figure 1. Gestational age-specific percentiles of mean arterial pressure (MAP) across pregnancy in the EDEN cohort. MAP percentiles for the full (N=1,875) and "low-risk", internal reference sample (N=795) are shown in panels A and B, respectively.**

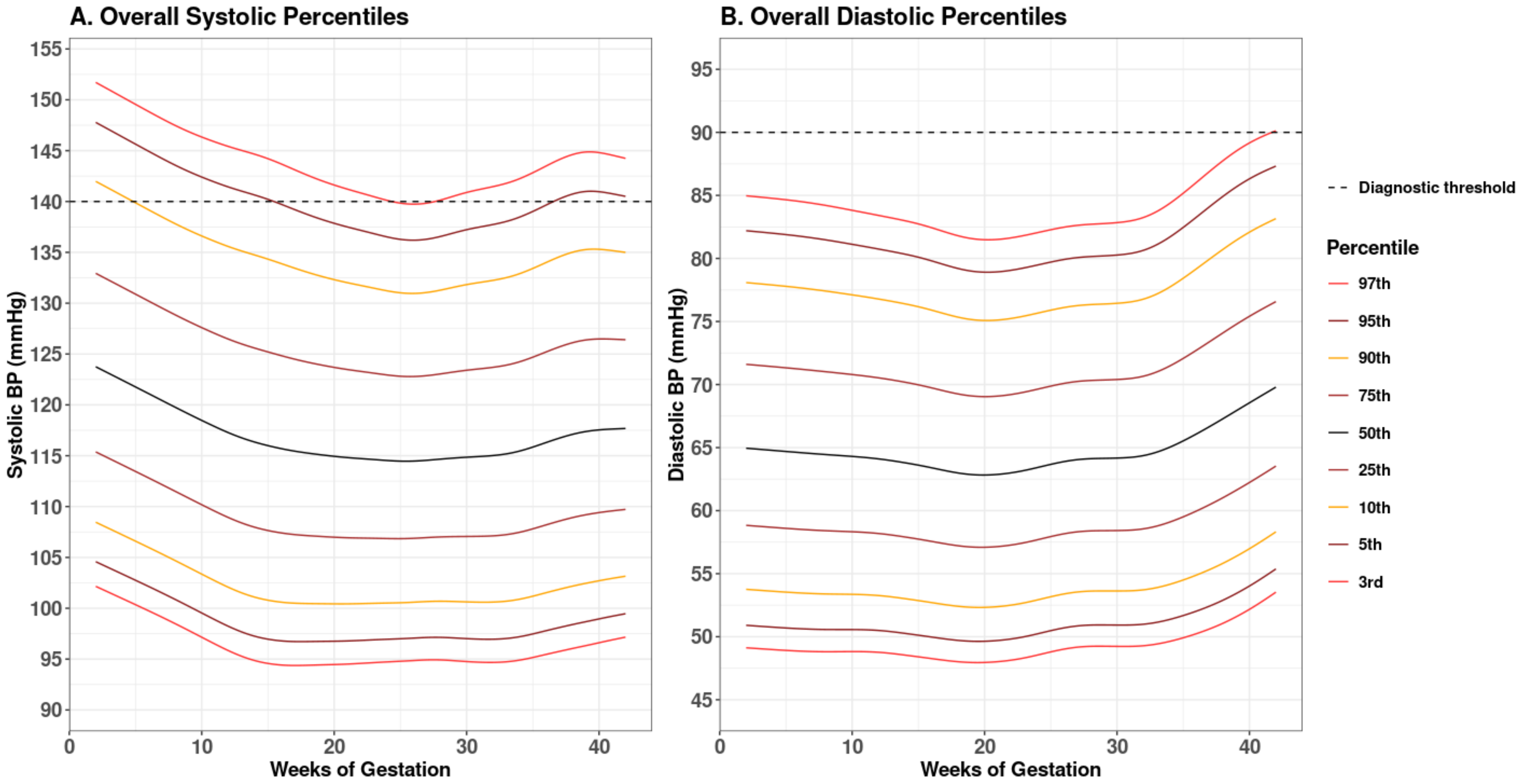


**Supplemental Figure 2. Overall, non-reference gestational age-specific percentiles of A. maternal systolic blood pressure (BP) and B. diastolic BP across pregnancy in the EDEN cohort, in the entire sample without exclusion (N=1875). These percentiles are not reference ranges, as they were defined in the overall sample and generated uniquely for descriptive purposes.**

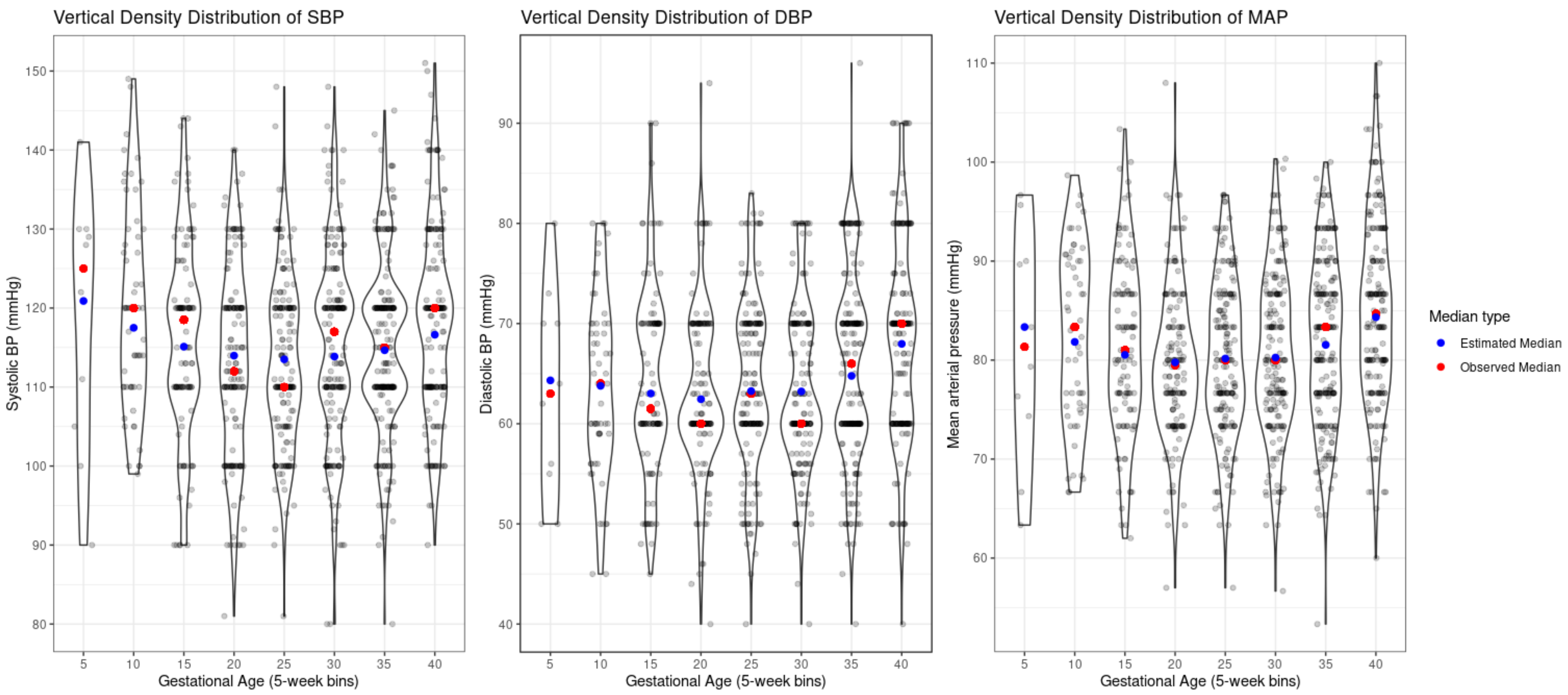


**Supplemental Figure 3. Internal empirical evaluation of gestational age-specific blood pressure (BP) reference percentiles in the EDEN cohort, overlaid on violin distribution plots of systolic BP (SBP), diastolic BP (DBP), and mean arterial pressure (MAP) distributions per 5-week gestational age window. Estimated median (50$^{th}$ percentile) values are shown in blue, and observed median values are shown in red.**

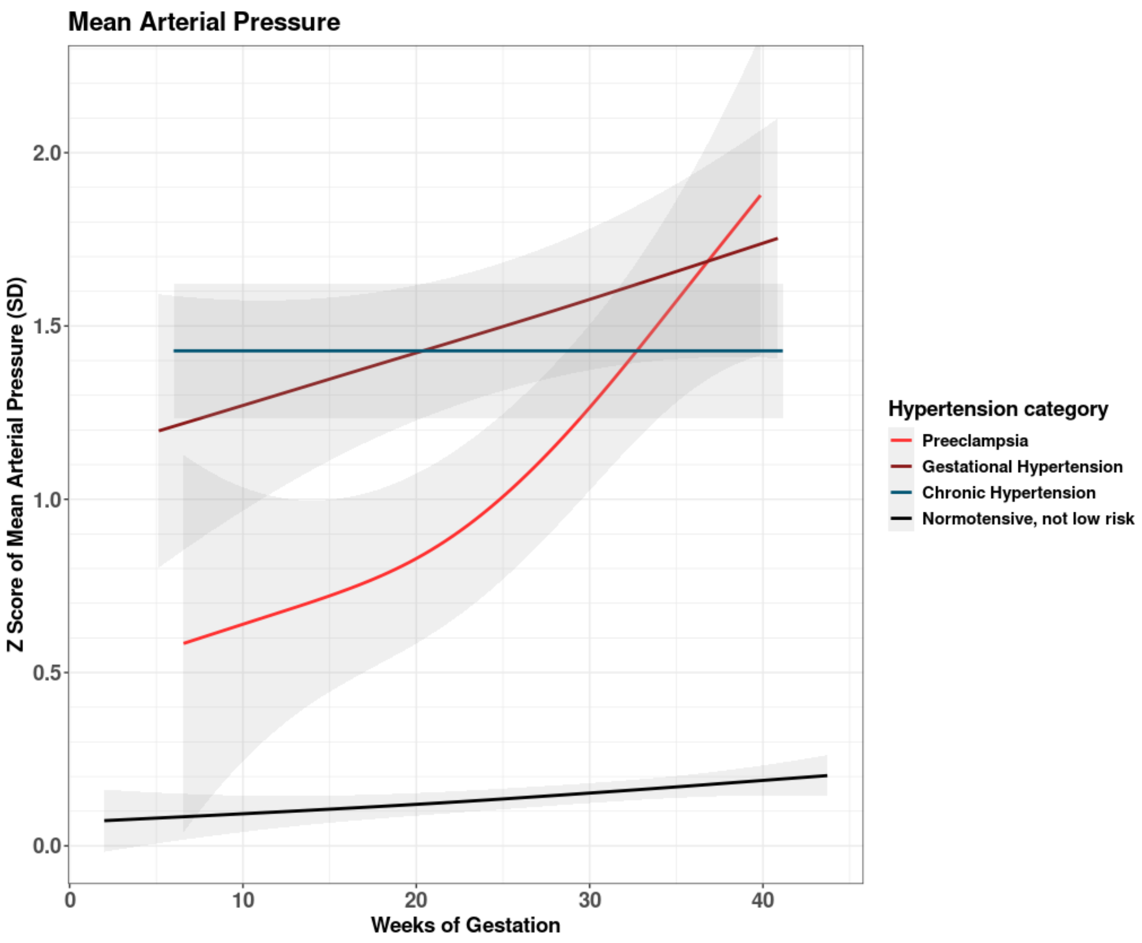


**Supplemental Figure 4. Gestational age-specific Z-scores of maternal blood pressure across pregnancy in the EDEN cohort, according to hypertensive disorders of pregnancy categories. Mean arterial pressure Z-score trajectories and 95% confidence intervals are shown. Data from normotensive women who were included in the "low-risk" reference subsample are excluded.**
