## Supplemental Methods for "Monitoring maternal blood pressure variations in pregnancy: gestational age-specific reference percentiles and Z-scores"

### Supplemental Methods (S1)

The GAMLSS framework allows for the estimation of four parameters: Mu (median), Sigma (variance), Nu (skewness), and Tau (kurtosis). We applied cubic splines (‘cs()’ in R), with 5 degrees of freedom for smoothing and used the ‘RS’ method for penalized maximum likelihood estimation. We tested multiple models, progressively increasing in complexity, and compared each model’s fit using global deviance (log-likelihood indicator) and worm and bucket plots.

Following comparisons using deviance measures and bucket plots, we selected models for systolic blood pressure (BP), diastolic BP, and mean arterial pressure (MAP) that 1) included both Mu and Sigma as functions of gestational age with cubic spline smoothers, 2) allowed Nu to vary without imposing a specific functional form, and 3) fixed Tau at its default value (for the BCPE distribution, this value is 2).

We chose to fix Tau for two reasons: first, to enhance the interpretability of the model, and second, to facilitate the generation of Z-scores using Lambda-Mu-Sigma (LMS) reference values derived from the model. Since our study aimed not only to create gestational age-specific percentile curves, but also to enable future applications of derived Z-scores, we opted against estimating Tau as a function of gestational age and allowing it to vary. While kurtosis (Tau) deviated from the ideal in the final models, it does improve when other parameters are estimated.

All model diagnostics are illustrated in Supplemental Methods Figures 1-5 using bucket plots, a type of diagnostic plot included in the GAMLSS package, which highlight residual errors related to skewness and kurtosis. Initial models are specified as M0, then as M1 and M2 when consequentially increasing in complexity. **Supplemental Methods Figures 1A-B** (systolic, M0 and M1) and **1C-D** (diastolic, M0 and M1) display fits for the "low-risk" internal reference sample. Notably, in this sample, allowing skewness (Nu) to vary partially accounted for kurtosis (Tau).

When further increasing complexity by modeling skewness (Nu) as a function of gestational age (**Supplemental Methods Figure 2A** (systolic, M2) and **2B** (diastolic, M2)), we did not observe substantial improvement in neither global deviance nor the bucket plots, therefore justifying our final model choice of M1 for both systolic and diastolic, in both the overall and restricted samples.

These same tendencies are illustrated in MAP, see: **Supplemental Methods Figures 3A-B** (M0 and M1, model without a function on skewness) and **3C** (Model with a function on skewness). Therefore, M1 was also chosen as the final model for MAP. Additionally, the bucket plots for this model of MAP demonstrated minimal estimation error for kurtosis, even though we did not specifically estimate kurtosis using model parameters.

**Supplemental Methods Figures: Diagnostic plots (next page)**

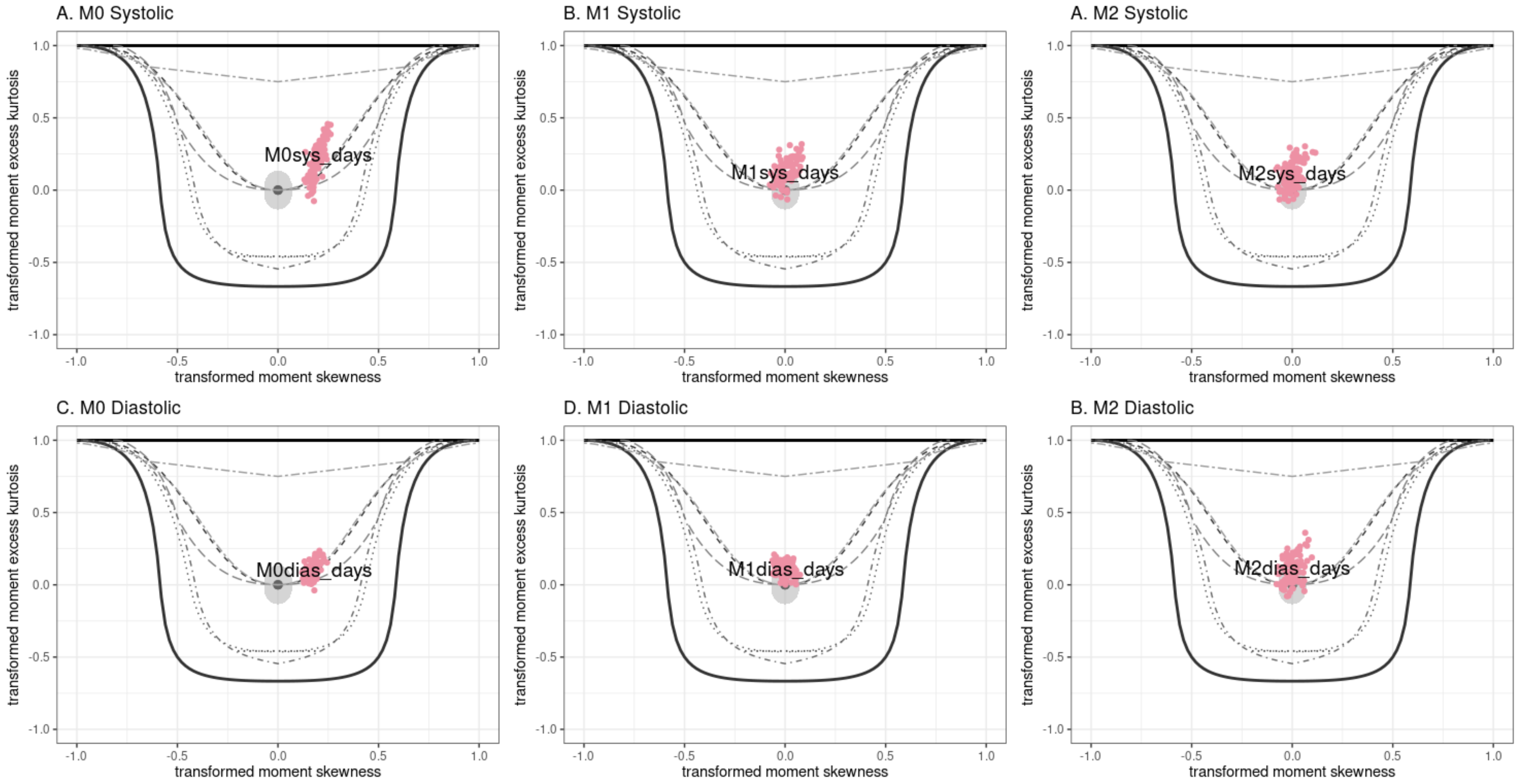


**Supplemental Methods Figure 1.** Bucket plots evaluating residual skewness and kurtosis estimation errors of the GAMLSS models for gestational age-specific references percentiles of systolic (A-B) and diastolic (C-D) blood pressure in the EDEN cohort. M0 = Mu and Sigma modeled as a function of gestational age; M1 = M0 + allow Nu to vary without imposing functional form; M2 = Mu, Sigma, and Nu modeled as a function of gestational age.

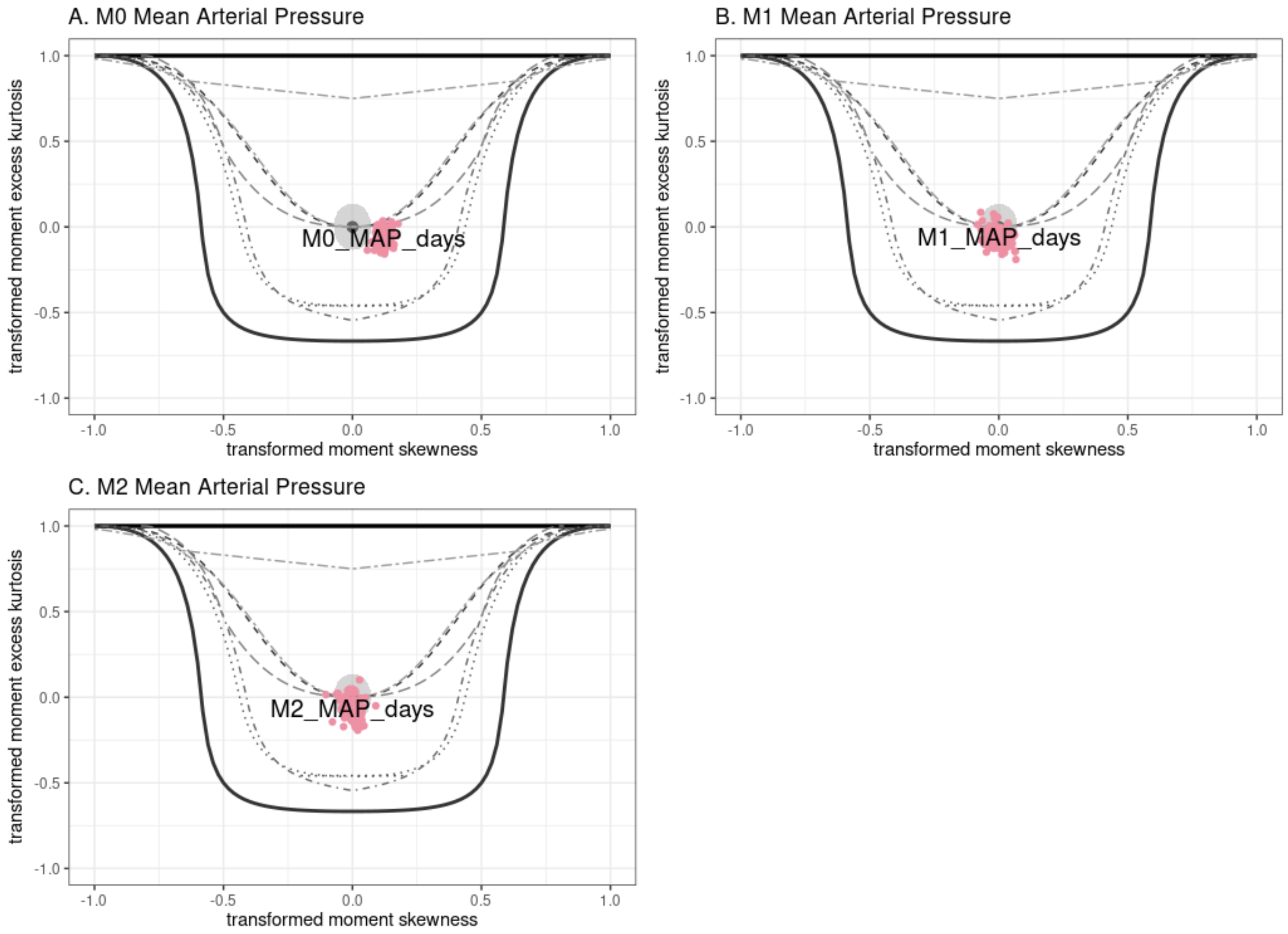


**Supplemental Methods Figure 2.** Bucket plots evaluating residual skewness and kurtosis estimation errors of the GAMLSS models for gestational age-specific references percentiles of mean arterial pressure in the EDEN cohort. M0 = Mu and Sigma modeled as a function of gestational age; M1 = M0 + allow Nu to vary without imposing functional form; M2 = Mu, Sigma, and Nu modeled as a function of gestational age.
