## Supplemental Table 1 for "Monitoring maternal blood pressure variations in pregnancy: gestational age-specific reference percentiles and Z-scores"

**Supplementary Table 1**

**Supplementary Table 1. Overall vs. “Low Risk” sample characteristics**

| ***Maternal characteristics*** | **Overall N = 1,875**[1] | **“Low-Risk” N = 795**[1] |
|---|---|---|
| **Age at delivery (years)** | 29 (26, 33) | 30 (27, 32) |
| *Missing* | *1* | *0* |
| **Country of birth** | | |
| Metropolitan France | 1,786 (95%) | 754 (96%) |
| French Departments / Territories | 13 (0.7%) | 3 (0.4%) |
| Other | 75 (4.0%) | 31 (3.9%) |
| *Missing* | *19* | *7* |
| **Education level** | | |
| Less than high school | 529 (28%) | 161 (20%) |
| High school diploma | 332 (18%) | 133 (17%) |
| 2-5 years of university studies | 999 (53%) | 498 (63%) |
| *Missing* | *15* | *3* |
| **Parity** | | |
| Primiparous | 833 (44%) | 357 (45%) |
| Multiparous | 1,039 (55%) | 438 (55%) |
| *Missing* | *3* | *0* |
| **Pre-Pregnancy BMI Status** | | |
| Normal weight: BMI < 25 $kg/m^2$ | 1,357 (74%) | 653 (82%) |
| Overweight: BMI [25-30) $kg/m^2$ | 324 (18%) | 142 (18%) |
| Obesity: BMI ≥ 30 $kg/m^2$ | 159 (8.6%) | - |
| *Missing* | *35* | *0* |

[1] Median (IQR); n (%)
